# Mutational pressure drives enhanced release of proteasome-generated public CD8^+^ T cell epitopes from SARS-CoV-2 RBD of Omicron and its current lineages

**DOI:** 10.1101/2024.04.03.24305074

**Authors:** Anna A. Kudriaeva, Ivan O. Butenko, George A. Saratov, Maxim Ri, Yuliana A. Mokrushina, Alexey A. Bondarev, Alena S. Evpak, Ivan V. Smirnov, Daria S. Matyushkina, Alexander G. Gabibov, Vadim M. Govorun, Alexey A. Belogurov

## Abstract

The COVID-19 pandemic was the most dramatic in the newest history with nearly 7 million deaths and global impact on mankind. Here we report binding index of 305 HLA class I molecules from 18,771 unique haplotypes of 28,104 individuals to 821 peptides experimentally observed from spike protein RBD of 5 main SARS-CoV-2 strains hydrolyzed by human proteasomes with constitutive and immune catalytic phenotypes. Our data read that mutations in the hACE2-binding region RBD_496-513_ of Omicron B.1.1.529 strain results in a dramatic increase of proteasome-mediated release of two public HLA class I epitopes. Global population analysis of HLA class I haplotypes, specific to these peptides, demonstrated decreased mortality of human populations enriched in these haplotypes from COVID-19 after but not before December, 2021, when Omicron became dominant SARS-CoV-2 strain. Noteworthy, currently circulating BA.2.86 and JN.1 lineages contain no amino acid substitutions in RBD_496-513_ thus preserving identified core epitopes.

## INTRODUCTION

Coronavirus disease 2019 (COVID-19) caused by SARS-CoV-2 infection is associated with immune dysregulation and abnormal hyperinflammation and on January 2024 reached 0.7 billion confirmed cases with 1% mortality (World Health Organization). Despite lost pandemic status, SARS-CoV-2 is still actively persisting in the human population. SARS-CoV-2-specific CD4+ and CD8+ T cell responses but not antibody titer play a critical protective role during COVID-19 [1]. Kinetic studies quantifying SARS-CoV-2-specific T cells and antibody responses over the course of acute infection in naive individuals revealed rapid expansion of IFNγ- producing T cells specific for different structural and non-structural SARS-CoV-2 proteins including spike (S), membrane (M), nucleocapsid protein (N), ORF3a and ORF7/8 in patients who experienced rapidly controlled SARS-CoV-2 replication without severe disease. In contrast, patients with prolonged infection and severe COVID-19 mounted robust antibody responses but had less circulating SARS-CoV-2-specific T cells [2]. Over 2,400 experimentally verified human T cell SARS-CoV-2 class I and class II epitopes have been described so far, and in majority of subjects T cell responses exhibit remarkable breadth [3,4].

According to World Health Organization there are 5 SARS-CoV-2 variants of concern (VOCs), which have been circulating: Alpha (B.1.1.7), Beta (B.1.351), Gamma (B.1.1.28(P.1)), Delta (B.1.617.2), and Omicron (B.1.1.529, including BA.1, BA.2, BA.3, BA.4, BA.5, and descendent lineages). At the present time no SARS-CoV-2 variants meet the VOC criteria, but several variants are stated as Variant of Interest (VOI) and Variants under monitoring (VUM) including the emerging lineage of Omicron BA.2.86 [5]. Current data indicate that the Omicron variant has the largest number of mutations, and thus the highest ability to evade therapeutic monoclonal and vaccine-elicited polyclonal neutralizing antibodies [6–10]. On the other hand, the T cell responses against the Omicron variant in donors, who were vaccinated with ancestral S protein or infected by previous strains, were largely preserved [11–14].

The intrinsic severity of Omicron is lower than that of previous variants, potentially due to its tropism for upper airways and hence lower risk of lower respiratory tract infection [15]. Mutations in Omicron spike protein as well as in nsp6 causes less severe disease compared to the ancestral virus due to altered viral tropism and adaptation to the changed tissue environment [16]. The less efficient spike cleavage of Omicron at S1/S2 [17] is associated with a shift in cellular virulence away from TMPRSS2-expressing cells, with implications for altered pathogenesis [18]. Additionally, Omicron variant is less effective in antagonizing the host cell interferon response, which may explain why it cause less severe disease [19].

Summarizing, despite SARS-CoV-2 lost its pandemic status, comprehensive analysis of its evolution and immunogenicity features may better prepare humanity to overcome more confidently future SARS-like viral epidemics. In our study we aimed to elucidate if there any antigenic determinants in Omicron variant, which may contribute to its decreased severity. Here we experimentally analyzed fragmentation of spike protein receptor binding domain (RBD) of five main SARS-CoV-2 strains by human proteasomes of constitutive and immune phenotypes. Our data revealed that reduced severity of Omicron variant may be directly linked with the mutations in its C-terminal region causing altered proteasome-mediated release of two CD8^+^ T cell-positive immunodominant HLA class I epitopes, covering 82% and 27% of world population haplotypes. Analysis of population frequency of HLA class I alleles revealed that *HLA-B*07:02*, - *B*08:01, -B*15:01, -C*01:02, -C*06:02* and *-C*07:02* potentially provides increased resistance of human population to Omicron.

## RESULTS

### Study design

Purified human 20S proteasomes from HeLa cells (constitutive proteasomes, c20S) and HeLa cells exposed to IFNγ (immunoproteasomes, i20S) (**Figure 1A**) were mixed with mammalian-expressed RBD variants of SARS-CoV-2 strains Wuhan-Hu-1, Alpha B.1.1.7, Delta B.1.617.2, Gamma B.1.1.28 (P.1) and Omicron B.1.1.529 (**Figure 1B**). Obtained hydrolysates were analyzed by liquid chromatography-tandem mass spectrometry (LS-MS/MS) (**Figure 1C**). Peptides from 9 to 16 amino acids long were subjected to *in silico* ERAAP (aminopeptidase associated with antigen processing in the ER [20,21]) N-terminal truncation to length of 8-10 amino acids [22] and analyzed versus HLA class I molecules representing 305 alleles of 18,771 unique haplotypes from 28,104 individuals according to Allele Frequency Net Database (98% world population dataset coverage) (**Figure 1D**). The HLA class I EL ranks calculated by artificial neural network netMHCpan-4.1 were used for further analysis of population haplotype HLA class I protective index, count of RBD-positive HLA class I haplotypes and estimation of SARS-CoV-2 mutation drift on its immunogenicity (**Figure 1E**).

**Figure 1.**
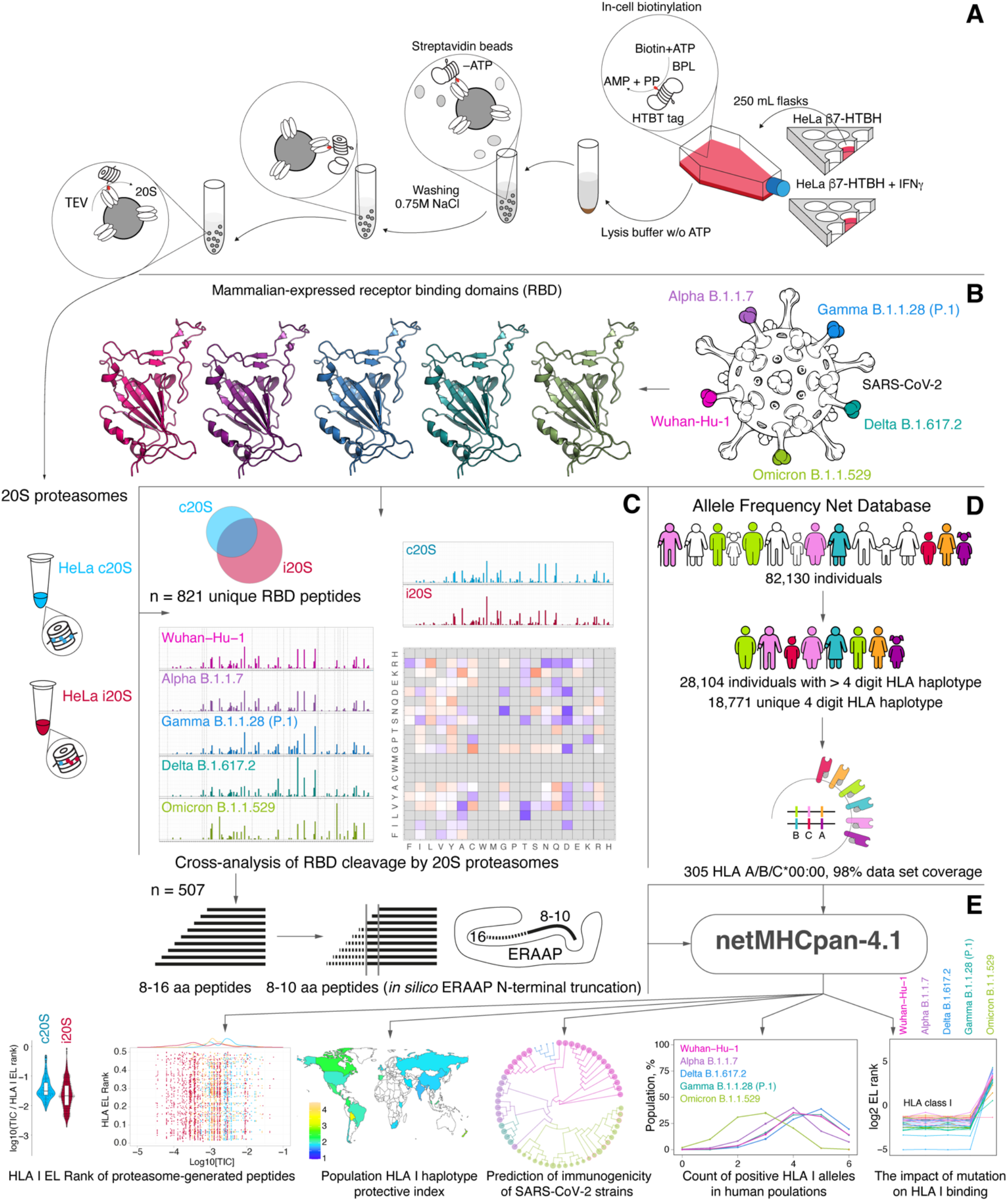
Study design. **(A)** Intact HeLa cells or HeLa cells treated by IFN𝛾 stably expressing HTBH-tagged proteasome subunit β7 were lysed in the absence ATP and further protein lysates were incubated with streptavidin beads. Washing buffer was supplemented with 0.75 M NaCl to dissociate 20S and 19S particles. Trapped human constitutive proteasomes (HeLa c20S) and immunoproteasomes (HeLa i20S) were eluted with TEV protease. Mammalian-expressed (**B**) receptor-binding domains (RBDs) of SARS-CoV-2 strains Wuhan-Hu-1, Alpha B.1.1.7, Delta B.1.617.2, Gamma B.1.1.28 (P.1) and Omicron B.1.1.529 were hydrolyzed by 20S proteasomes with different catalytic phenotypes and further analyzed by LS-MS/MS (**C**). Observed 8-16 amino acid peptides (n = 507) were *in silico* truncated to length of 8-10 and further analyzed by netMHCpan-4.1 algorithm for binding index with 305 HLA class I molecules covering 18,771 unique haplotypes of 28,104 individuals from Allele Frequency Net Database (**D**).(**E**) Final data included EL rank values of proteasome-generated peptides, population haplotype HLA class I binding index, and count of RBD-positive HLA class I haplotypes.

There are several reasons why we chosen uncapped 20S proteasome for our *in vitro* studies. Firstly, 20S proteasomes are believed to represent two-thirds of the total proteasome pool [23] and may degrade up to 20% of intracellular proteins [24]. At least 10% of HLA class I peptides were found to be dependent on the proteasome but independent of ubiquitination for their generation [25]. Secondly, 20S proteasomes itself may degrade ubiquitinated substrates [26], especially in oxidative stress conditions [27,28]. Thirdly, SARS-CoV-2 S protein ubiquitination [29] is post-translational modification, which affects its cleavage [30] rather is linked with ubiquitin-dependent degradation mediated by 26S proteasome.

### Fragmentation of RBD variants from 5 reference SARS-CoV-2 strains by human proteasomes with different catalytic phenotypes

The DNA constructs coding for HTBH-tagged proteasome subunit β7 (PSMB4) were integrated into the genome of the HEK 293T and HeLa cells using Sleeping Beauty transposon system as described previously [31]. HEK 293T and HeLa cells, exposed or not exposed to IFN𝛾 for 96 hours, stably expressing HTBH-tagged proteasome subunit, were lysed in the absence ATP and further protein lysates were incubated with streptavidin beads. Washing buffer was supplemented with 0.75 M NaCl to dissociate 20S and 19S particles. Purified human proteasomes were analyzed by gel electrophoresis utilizing Coomassie staining, in-gel fluorescence with Me_4_BodipyFL-Ahx_3_Leu_3_VS fluorescent proteasome probe (UbiQ18) [32] (**Figure 2A**) and western blotting (**Figure 2B**). Our data clearly indicate that in contrast to HeLa and HEK 293T cells, 20S proteasome isolated from HeLa cells subjected to IFNγ has distinct immune phonotype due to the presence of all three catalytic subunits β1i, β2i and β5i. In line with this, activity toward immunoproteasome substrate PAL-AMC and ratio of chymotryptic to caspase activity of 20S proteasomes, isolated from HeLa cells exposed to IFN𝛾, increased approximately 2.5 times in comparison with non-treated HeLa and HEK 293T cells (**Figure 2C**).

**Figure 2.**
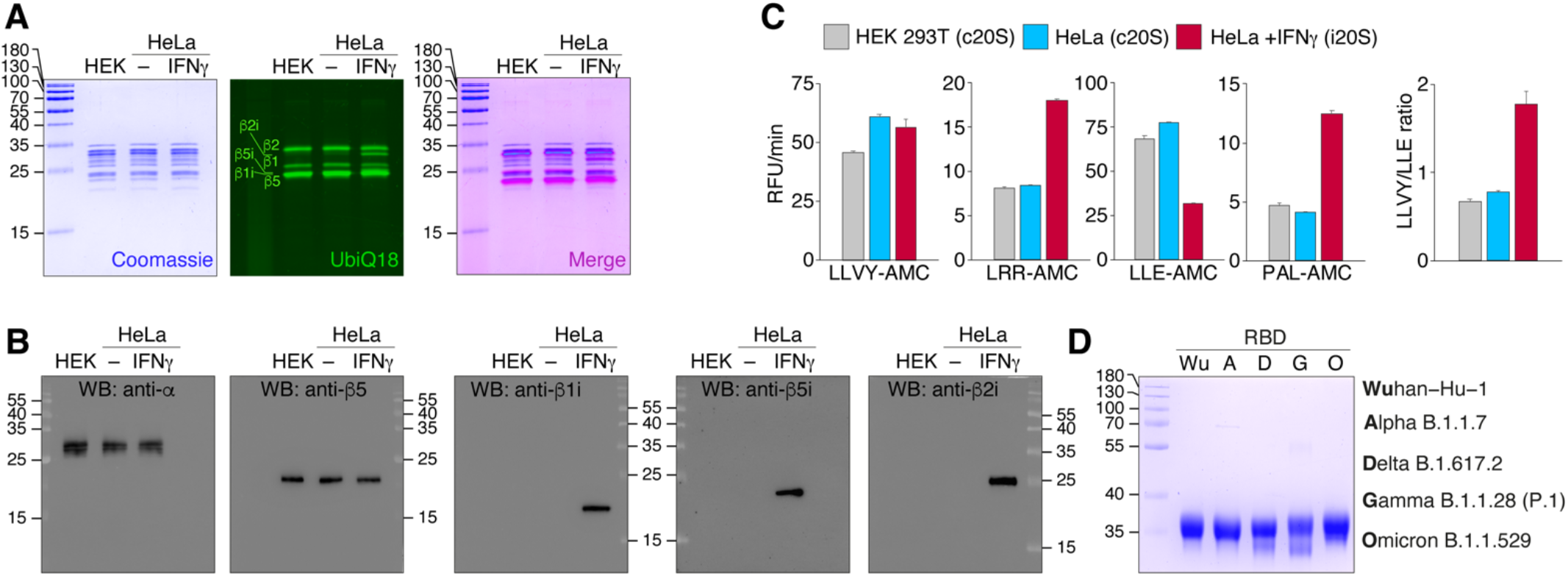
Characterization of purified human c20S and i20S proteasomes from HEK 293T and HeLa cells. Analysis of the proteasomes purified from HEK 293T, HeLa cells and HeLa cells treated by IFN𝛾 by Coomassie-stained SDS-PAGE and in-gel fluorescence using Me4BodipyFL-Ahx3Leu3VS fluorescent probe (UbiQ18) (**A**), western blotting with antibodies specific to structural (α) and catalytic (β) core subunits (**B**). (**C**) Activity of purified proteasomes toward fluorogenic 7-amido-4-methylcoumarin (AMC) substrates. The LLVY-AMC/LLE-AMC activity ratio is shown right. (**D**) The S protein RBDs purified from HEK 293F cells of Wuhan-Hu-1, Alpha B.1.1.7, Delta B.1.617.2, Gamma B.1.1.28 (P.1) and Omicron B.1.1.529 strains analyzed by SDS-PAGE followed by Coomassie staining.

Due to the identical subunit composition and activity of 20S proteasomes, isolated from HEK 293T cells and HeLa cells not exposed to IFN𝛾, in further experiments we used constitutive 20S proteasome (c20S) from untreated HeLa cells and immunoproteasome-enriched 20S samples from HeLa cells subjected to IFN𝛾 (i20S). The S protein RBDs purified from HEK 293F cells of Wuhan-Hu-1, Alpha B.1.1.7, Delta B.1.617.2, Gamma B.1.1.28 (P.1) and Omicron B.1.1.529 strains (**Figure 2D**) were incompletely (80-90%) digested by c20S and i20S proteasomes in order to restrict substrate re-entry. The LS-MS/MS analysis of hydrolysates revealed different distribution of C-terminal and N-terminal amino acids in peptides, generated by c20S and i20S proteasomes (**Figure 3A**). Both types of proteasomes tended to generate peptides with hydrophilic N-terminus, whereas C-terminus of i20S-released RBD peptides were more hydrophobic in comparison with c20S proteasomes. As expected, c20S proteasomes more actively hydrolyze RBD variants after acidic amino acids, especially aspartate, due to the presence of caspase-like activity. Analysis of sequence-specific RBD cleavage between c20S and i20S revealed that difference in cleavage sites between c20S and i20S was similar between RBD variants and was minimal in Omicron B.1.1.529 RBD variant (**Figure 3B**).

**Figure 3.**
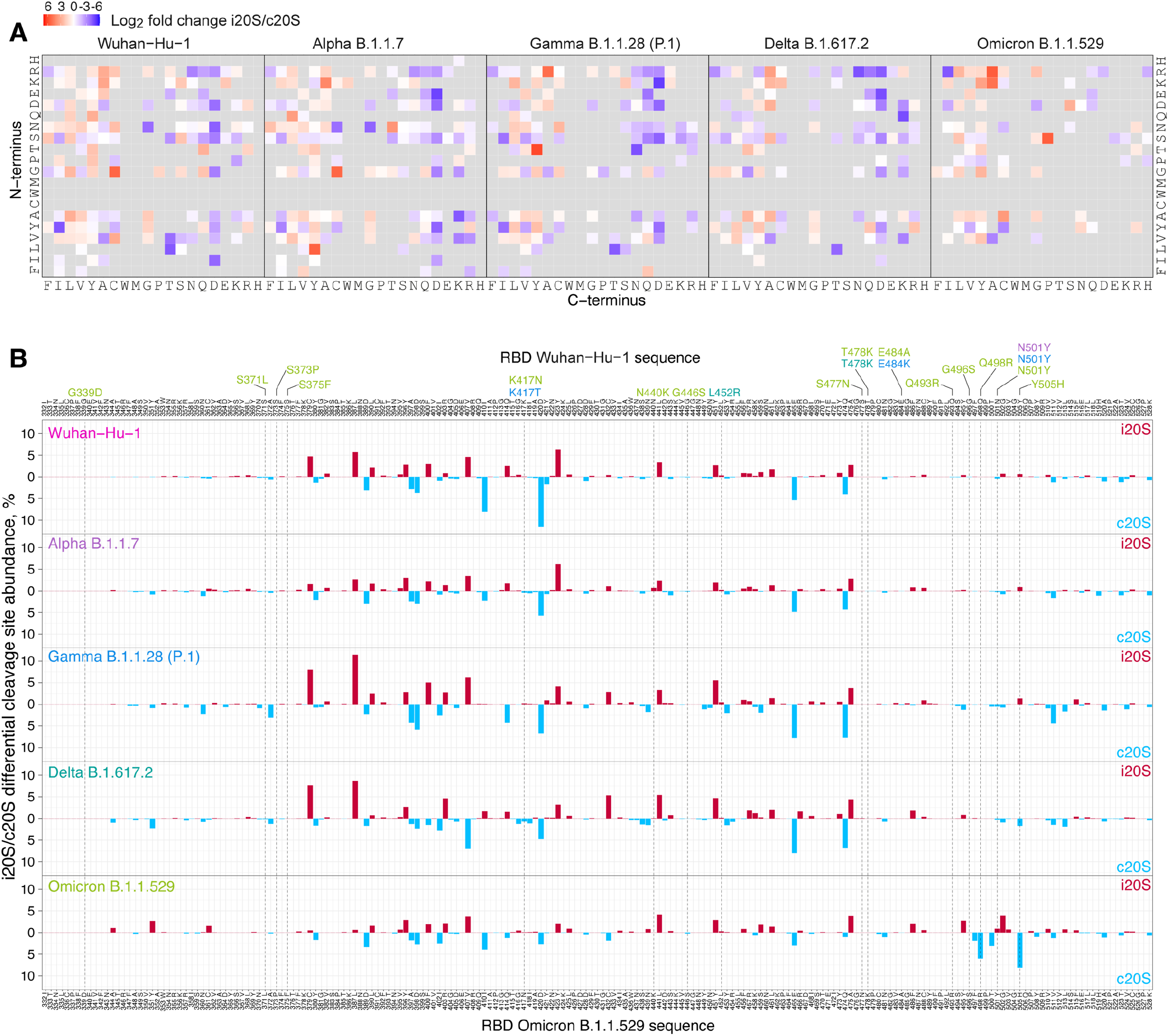
The SARS-CoV-2 S protein RDB cleavage site preferences of human proteasomes with different catalytic phenotypes. (**A**) Differential plot of peptides N- and C-terminal amino acids from RBD variants hydrolyzed by human proteasomes with different catalytic phenotypes (c20S and i20S). (**B**) Differential abundance of c20S (blue) and i20S (red) cleavage sites in RBD of SARS-CoV-2 strains Wuhan-Hu-1 (pink), Alpha B.1.1.7 (violet), Gamma B.1.1.28 (P.1) (blue) Delta B.1.617.2 (aquamarine) and Omicron B.1.1.529 (light green). Mutations in RBD sequence are indicated by respective colors.

The individual peptide patterns (**Figure 4A**) revealed that i20S proteasomes generated from RBD variants the highest diversity of peptides with length more than 7 amino acids, except Omicron RBD. The c20S-generated RBD peptides were significantly less variable and mostly overlapped with peptides produced by immunoproteasomes i20S. Total amount of detected peptides was minimal in Omicron B.1.1.529 samples. Proteasome-mediated RBD cleavage was the most intensive in the middle part of the protein, and flanking regions approximately 50 amino acids long were significantly less processed. Distribution of major sites of RBD cleavage demonstrated similar pattern of hydrolysis of RBD from Wuhan-Hu-1, Alpha B.1.1.7, Delta B.1.617.2, Gamma B.1.1.28 (P.1) strains (**Figure 4B**), whereas Omicron B.1.1.529 RBD variant was significantly higher processed in the 495-505 region in comparison with other strains.

**Figure 4.**
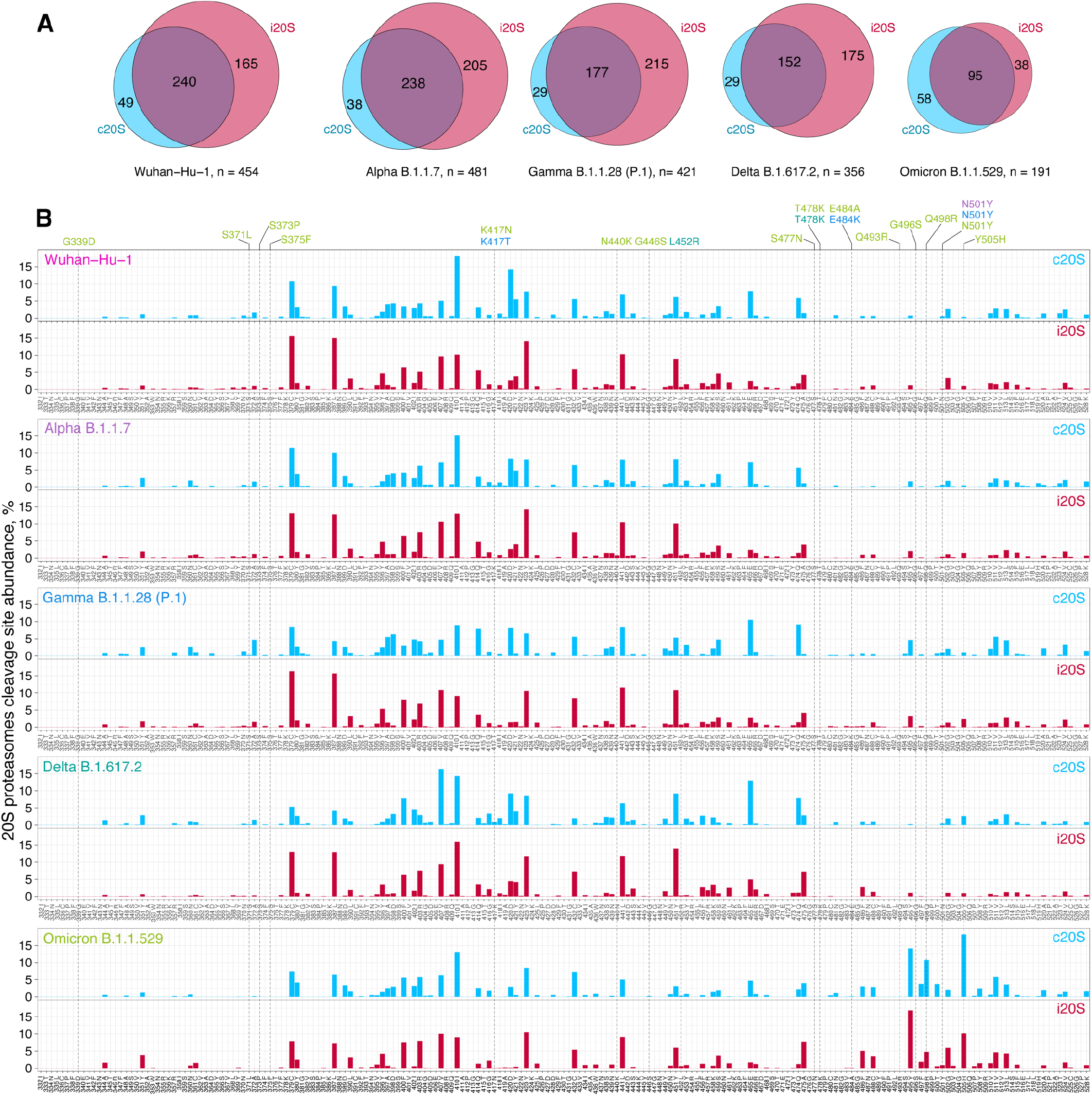
Fragmentation of SARS-CoV-2 S protein RBD variants by human proteasomes with different catalytic phenotypes. (**A**) Venn diagrams representing distribution of RDB peptides after proteasome-mediated (i20S proteasomes in blue, i20S – in red) hydrolysis identified by LS-MS/MS. (**B**) Relative intensity of RBD cleavage sites of SARS-CoV-2 strains Wuhan-Hu-1 (pink), Alpha B.1.1.7 (violet), Gamma B.1.1.28 (P.1) (blue) Delta B.1.617.2 (aquamarine) and Omicron B.1.1.529 (light green). Mutations in RBD sequence are indicated by respective colors.

### Prediction of affinity of S protein RBD HLA class I epitopes utilizing LC-MS/MS data of RBD hydrolysis by proteasomes with different catalytic phenotypes

Experimentally observed peptides from 8 to 16 amino acids long were further subjected to *in silico* ERAAP N-terminal truncation to length of 8-10 amino acids and analyzed versus HLA class I molecules corresponded to 305 alleles (**Supplemental Tables 1, 2 and 3, Supplemental Data 1 and 2**). Analysis of distribution of peptides with EL rank less than 0.5 revealed that i20S immunoproteasomes solely generate approximately half of total HLA I epitopes, whereas other part is mostly generated by both, c20S and i20S (**Figure 5A**). Percentage of peptides overlapping between c20S and i20S RBD hydrolysis products increased in Omicron B.1.1.529 RBD samples from 50 to 65 in comparison with other RBD variants. Analysis of HLA class I EL rank of the proteasome-generated RBD peptides versus its relative abundance (**Figure 5B**) revealed that HLA class I epitopes in Omicron B.1.1.529 RBD samples shifted to the more quantitively representative area. Plotting of density of HLA class I epitopes relatively its intensity clearly demonstrated that, firstly, i20S proteasomes generally produce broader repertoire of the peptides and, secondly, c20S generates HLA class I epitopes more intensively (**Figure 5C**). Importantly, in contrast to other strains, in the Omicron B.1.1.529 RBD samples c20S and i20S HLA class I epitopes density curves were completely overlapped. Calculation of median and interquartile range of relative abundance and its relative abundance to EL rank index of c20S- and i20S-generated HLA I epitopes revealed statistically significant increase of Omicron B.1.1.529 values in comparison with other SARS-CoV-2 RBD variants (**Figure 5D**). Finally, estimation of cumulative protection index of unique RBD HLA I epitopes for each variant demonstrated the highest values for Omicron B.1.1.529 RBD peptides generated by both, c20S and i20S proteasomes (**Figure 5E, Supplemental Table 4**).

**Figure 5.**
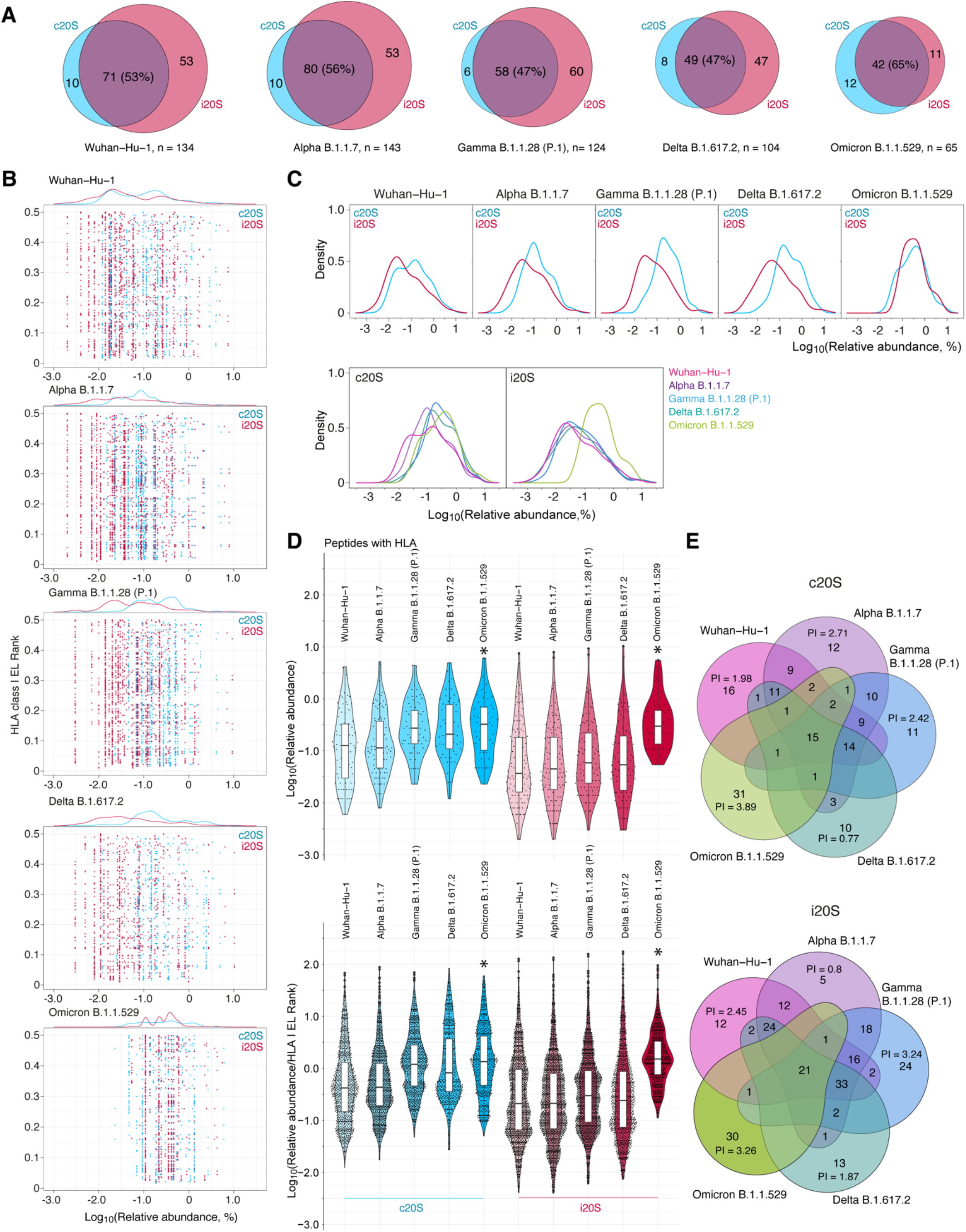
Analysis of HLA class I binding index of SARS-CoV-2 RDB peptides generated by human proteasomes with different catalytic phenotypes. **(A)** Venn diagrams representing distribution of proteasome-generated RDB peptides (herein and after c20S in blue, i20S – in red) with HLA class I EL rank ! 0.5. (**B**) Distribution of HLA class I EL rank and relative abundance of proteasome-generated RDB peptides related to different SARS-CoV-2 strains. (**C**) Density of HLA class I epitopes count (without duplication) grouped by SARS-CoV-2 strains (top) and proteasomes type (bottom). (**D**) Violin plots of HLA class I proteasome-generated peptide binders according to its relative abundance (top) and relative abundance normalized to HLA class I EL rank (bottom). Statistically significant difference with ancestorial Wuhan-Hu-1 strain is indicated by asterisk. (**E**) Multiple Venn diagrams of S protein RBD peptides of SARS-CoV-2 strains Wuhan-Hu-1 (pink), Alpha B.1.1.7 (violet), Gamma B.1.1.28 (P.1) (blue), Delta B.1.617.2 (aquamarine) and Omicron B.1.1.529 (light green) generated by c20S (top) and i20S (bottom) proteasomes. Protective index (average amount of epitope-positive HLA class I alleles in world population human haplotypes, PI) for unique RBD HLA class I epitopes is indicated.

### Elucidation of physiologically relevant SARS-CoV-2 S protein public RBD epitopes

Firstly, we collapsed proteasome-generated N-terminally extended peptides related to the same core HLA class I CD8^+^ T cell epitopes listed in the Immune Epitope Database (IEDB) (**Supplemental Tables 5 and 6**). Comparison of previously reported CD8^+^ T cell epitopes [3,4,33–40] and experimentally observed proteasome-generated SARS-CoV-2 core RBD HLA class I epitopes revealed that only minor part of them is overlapped (**Figure 6A, Supplemental Table 7**). Majority of abundant proteasome-generated epitopes have neither broad HLA class I haplotype coverage nor experimentally registered CD8^+^ T cell response. We further focused on two RBD regions, namely 496-513 and 442-453. In the C-terminal Omicron RBD region 496-513 we observed enhanced release of two core epitopes, namely _504_GHQPYRVVVL_513_ and _496_SFRPTYGVGH_505_ (**Figure 6B**). Epitope _504_GHQPYRVVVL_513_ was released from Omicron B.1.1.529 RBD variant by proteasome-mediated hydrolysis 3-10 times more effectively in comparison with _504_G**Y**QPYRVVVL_513_ in other SARS-CoV-2 RBD variants. Epitope _496_SFRPTYGVGH_505_ was observed solely in Omicron B.1.1.529 RBD hydrolysates (**Figure 6C**). Next, we systematically analyzed epitopes shown on Figure 6A in terms of c20S- and i20S proteasome-mediated release, percentage of HLA class I positive haplotypes (**Figure 6D**) and previously reported CD8^+^ T cell assays (**Figure 6E)**. Our data read that abundant proteasome-generated Omicron RBD HLA class I epitopes _504_GHQPYRVVVL_513_ and _496_SFRPTYGVGH_505_ cover 82 and 27 percentage of human haplotype variants and RBD_504-513_ elicited CD8 T cell response in 31% of individuals. Another representative Omicron RBD epitope _442_KVSGNYNYLY_451_ covers 17% of human haplotype variants and previously was shown to be positive in half of patients’ cytotoxic assays. In other SARS-CoV-2 RBD variants we highlighted RBD epitopes _506_QPYRVVVL_513_, _502_GVGYQPYRVV_511_ and less abundant _504_GYQPYRVVVL_513_, which were positive in one third of CD8^+^ T cell assays and cover 16, 35 and 97 percentage of human haplotypes variants, respectively.

**Figure 6.**
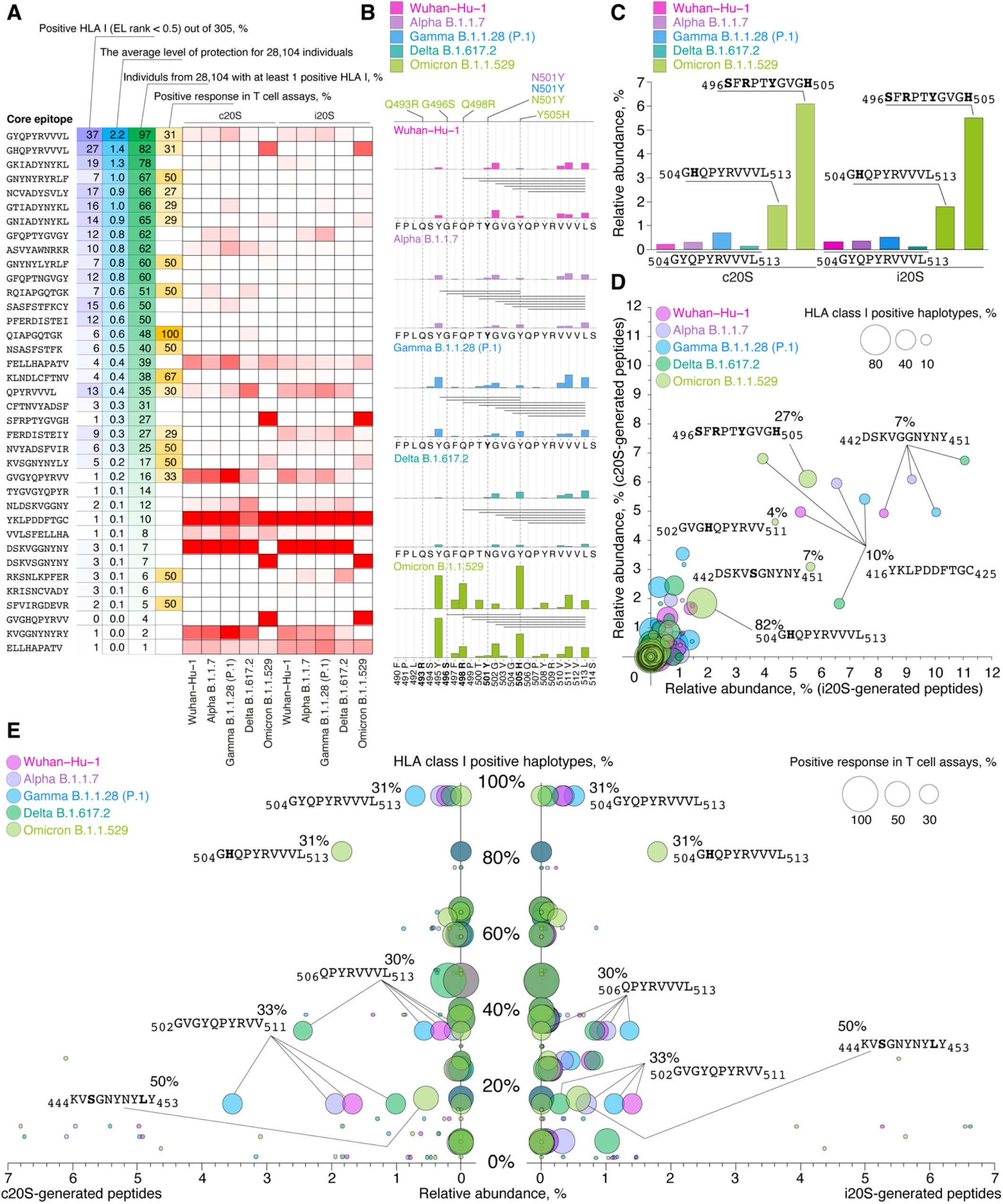
Proteasome-mediated Omicron RBD cleavage results in release of public CD8^+^ T cell core epitopes. (**A**) Comparison of previously reported CD8^+^ T cell epitopes and experimentally observed proteasome- generated collapsed core SARS-CoV-2 RBD HLA class I epitopes. (**B**) C-terminal Omicron RBD region 490-514 with indicated proteasome-generated N-terminally extended peptides and intensity of cleavage sites. (**C**) Relative abundance of core epitopes in RBD hydrolysates by c20S and i20S proteasomes. (**D,E**) Systematical analysis of epitopes shown on panel A in terms of c20S- and i20S proteasome-mediated release, percentage of HLA class I positive haplotypes (**D**) and previously reported CD8^+^ T cell assays (**E)**. Omicron mutations are shown in bold.

We further turned to the structural aspects of the determined main Omicron RBD HLA class I epitopes. To this aim we mapped these epitopes on previously reported [41] crystal structure of Omicron RBD-hACE2 complex (**Figure 7A**). Interestingly, amino acids from two of 3 epitopes, _496_SFRPTYGVGH_505_ and _442_DSKVSGNYNYLY_453_, are directly involved in the formation of the patch 2 of the RBD-hACE2 binding interface. Thus, Omicron B.1.1.529 RBD Y449 interacts with residues D38 and Q42 from hACE2, RBD R498 forms a salt bridge with D38 from hACE2, RDB T500 and Y501 interacts with Y41 from hACE2, and K353 from hACE2 forms a hydrogen bond with G502 of RBD [41].

**Figure 7.**
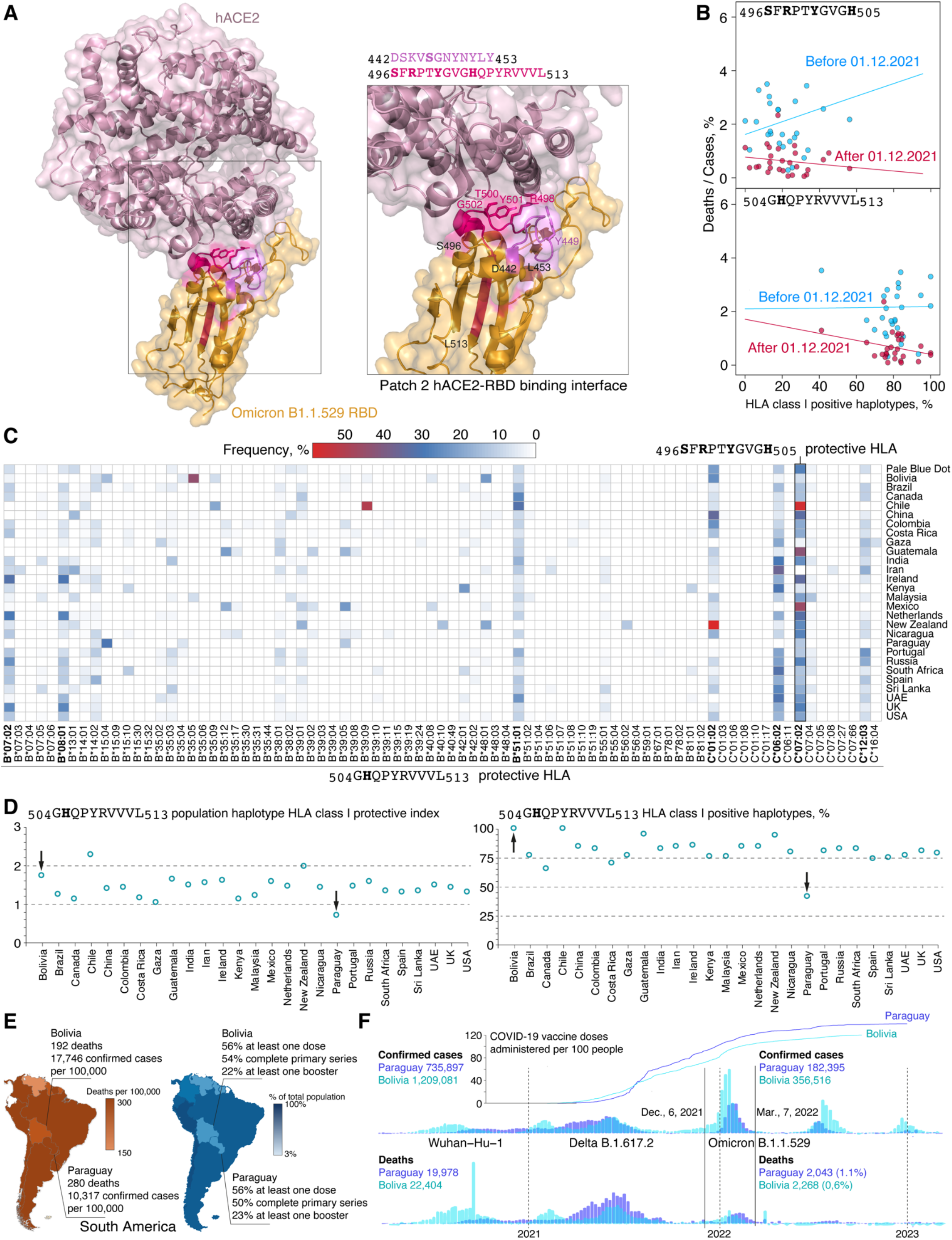
Identified proteasome-generated public Omicron SARS-CoV-2 RBD epitopes have global physiological relevance. **(A)** Crystal structure of Omicron RBD-hACE2 complex with marked regions containing public Omicron SARS-CoV-2 RBD epitopes. RBD amino acids crucial for patch 2 of the RBD-hACE2 binding interface are shown. (**B,C**) Correlation between COVID-19 mortality and percentage of haplotypes (**B**) and population frequency of haplotypes (**C**) positive toward public Omicron SARS-CoV-2 RBD epitopes in different countries. Omicron mutations are shown in bold. (**D**) Analysis of population haplotype protective index and HLA class I positive haplotypes frequency toward RBD503-513 core epitope in different countries. (**E**) COVID-19 mortality rate and vaccination status in Republic of Paraguay and geographically proximal Plurinational State of Bolivia in South America. (**F**) Mortality rate, confirmed COVID-19 cases and vaccination dynamics in Paraguay and Bolivia since March, 2020 till beginning of 2023

Global population analysis of HLA class I haplotypes specific to Omicron RBD peptides _496_SFRPTYGVGH_505_ and _504_GHQPYRVVVL_513_ demonstrated negative correlation between COVID-19 mortality and percentage of positive haplotypes in distinct countries (**Supplemental Tables 8 and 9**). Remarkably, this correlation was observed after but not before December, 2021, when Omicron B.1.1.529 spread over the world and became dominant SARS-CoV-2 strain (**Figure 7B**). Analysis of population frequency of _496_SFRPTYGVGH_505_ and _504_GHQPYRVVVL_513_-binding HLA class I molecules revealed that *HLA-B*07:02, -B*08:01, - B*51:01, -C*01:02, -C*06:02 and -C*07:02* potentially may provide increased resistance of human population to Omicron B.1.1.529 strain (**Figure 7C**).

Evaluation of population-specific HLA class I haplotypes, which may present Omicron RBD _504_GHQPYRVVVL_513_ peptide revealed abnormally low protective index (average count of binding alleles) and percentage of positive haplotypes (haplotypes with at least one binding allele) in Republic of Paraguay, South America (**Figure 7D**). We next analyzed COVID-19 mortality rate and vaccination status in Paraguay and geographically proximal Plurinational State of Bolivia (**Figure 7E**). According to the World Health Organization, value of deaths caused by COVID-19 was 1.5 times lower in Bolivia than in Paraguay. Herewith, frequency of confirmed SARS-CoV-2 infection cases was significantly higher in Bolivia in comparison with Paraguay, whereas current vaccination status in both countries is similar. We further used World Health Organization weekly reports in order to track COVID-19 confirmed cases and deaths in Bolivia and Paraguay starting from March, 2020 till beginning of 2023. These data strikingly indicate almost doubled mortality rate per confirmed COVID-19 cases in Paraguay versus Bolivia during the fourth Omicron B.1.1.529-raised COVID-19 pandemic wave (**Figure 7F**). At the same time, dynamic of vaccination per 100 people in Bolivia and Paraguay was nearly identical.

## DISCUSSION

Despite numerous studies describing potent SARS-CoV-2 HLA class I-binding peptides and T cell epitopes, absolute majority of these studies operate with T cell assays using overlapping synthetic peptides or *in silico* prediction algorithms [42,43]. As a result, proteasome, which generates majority of HLA class I-exposed peptides, is beyond the resulting conclusions. Consequently, putting proteasome-mediated antigen processing out of the brackets may significantly misrepresent interpretation of such datasets as reported epitopes may neither exist [44] nor has any physiological meaning. Another crucial aspect is that current knowledge on SARS-CoV-2 CD8^+^ T cell epitopes evidently are restricted by most frequent human haplotypes. Here we aimed to cover this gap by comprehensive analysis of SARS-CoV-2 RBD immunopeptidome of 5 main SARS-CoV-2 strains released by human proteasomes with constitutive and immune phenotypes. As a result, we report binding index of 305 HLA class I molecules from 18,771 unique haplotypes of 28,104 individuals to 821 physiologically relevant RBD peptides.

Our data suggest that immunoproteasomes generate broader peptide repertoire, however, surprisingly it is less immunogenic in terms of integral HLA class I epitope quantity. Nonetheless, convolution of the relative abundance of N-terminally extended epitopes with identical C-terminus leads to nearly similar release of core CD8^+^ T cell epitopes by both types of proteasomes, which correlates with study by Cascio et al. [45]. In case of Omicron RBD constitutive and immune proteasomes generate less diversity but significantly more quantity of HLA class I epitopes in comparison with other variants. Middle part of RBD is processed more intensively, although, another possibility is that flanking fragments are, contrary, overprocessed and hydrolysis products are less than 6-8 amino acids in length [46]. Constitutive and immune proteasomes have distinct cleavage site preferences and this difference is less manifested in Omicron RBD variant. Also, proteasome-mediated hydrolysis of Omicron RBD variant results in diminished amount of peptides with length more than 8 amino acids, which may be caused by decreased stability and increased susceptibility to protease digestion of this variant [47]. Similar to flanking regions of all variants, Omicron RBD peptides may be overprocessed by both proteasomes in general.

Our data suggest that majority of previously reported CD8^+^ T cell SARS-CoV-2 RBD epitopes are not overlap with proteasome-generated peptides. Complex analysis of observed CD8 T cell epitopes, global HLA class I haplotypes frequency and relative abundance of the RBD peptides generated by proteasomes revealed two RBD regions, 496-513 and 442-453, which contains the most pronounced CD8^+^ T cell antigenic determinants. Quantity of three core RBD epitopes _504_GHQPYRVVVL_513_, _496_SFRPTYGVGH_505_ and _442_KVSGNYNYLY_451_, which may be presented by HLA class I molecules encoded by 80, 30 and 17 percent of human haplotypes, respectively, was significantly increased in Omicron hydrolysates in comparison with other RBD variants. Importantly, RBD peptide _506_QPYRVVVL_513_ had the highest magnitude of response in AIM assay among tested CD8^+^ T cell S protein-derived antigens [34]. Also, our data are in line with recent report suggesting that SARS-CoV-2 S protein RBD_484-508_ peptide elicits T cell IFNγ response in cells from naïve-to-infection and unvaccinated subjects with close contact with SARS-CoV-2-positive patients comparable to those observed in cells infected by SARS-CoV-2 pseudovirus [48].

Analysis of human haplotypes revealed that molecules encoded by frequently distributed alleles *HLA-B*07:02*, -*B*08:01, -B*51:01, -C*01:02, -C*06:02 and -C*07:02* may present RBD_496- 513_-derived epitopes. These data are correlated with observation made by Olafsdottir et al. concluding that S1-reactive CD8^+^ T-cell responses are strongly associated with presence of *HLA-C*07:02* and *HLA-B*07:02* alleles [49]. Global analysis of COVID-19 mortality rate in 27 countries and pairwise comparison of Bolivia-Paraguay single case provide statistically significant evidences that haplotypes coding for HLA class I molecules binding _496_SFRPTYGVGH_505_ and _504_GHQPYRVVVL_513_ may be regarded as protective against Omicron SARS-CoV-2. Evolution of SARS-CoV-2 during last year resulted in appearance of BA.2.86 lineage [50], first identified in August 2023, and its more recent and currently circulating descendant, JN.1 [51]. While data related to JN.1 are yet limited, SARS-CoV-2 BA.2.86 elicits CD8^+^ T cell responses similar to other Omicron lineages [52–54]. Noteworthy, BA.2.86 and JN.1 are massively mutated in RBD region 442-453 but contain no amino acid substitutions in fragment 496-513 [51]. Thus, at least two major epitopes identified in our study are retained in these SARS-CoV-2 variants.

Question, which is still enigmatic, if there is a link between positioning of epitopes in RBD structure and its immunological relevance? The N-terminal part of RBD_496-513_ contains amino acid residues essential for hACE2 binding [41] and key sites for the host adaptation of SARS-CoV-2 [55]. C-terminal part forms a β-strand required for the maintaining of overall RBD structure (**Figure 7A**). This structural feature probably restricts mutational drift of this part as all mutations are located in the N-terminal segment. We further suggest that RBD_496-513_ region probably was maturated in Omicron lineages as major immunodominant epitope under synergetic evolution pressure driven by gaining ability of cross-species transmission [56,57] and by beneficial proteasome-mediated processing resulted in more effective SARS-CoV-2 recognition by human immune system. Another possibility may be permanent virus persisting in chronically infected individuals [58] until immune system obtain ability to identify viral peptides in the HLA class I context. Mutations in Omicron RBD_496-513_ preserve public C-terminal core CD8^+^ T cell epitope and at the same time allow proteasome to generate increased amount of N-terminally extended pre-epitopes suitable for ERAAP truncation. Resulted decreased severity and asymptomatic SARS-CoV-2 infection evidently increase rate of human-to-human Omicron SARS-CoV-2 transmission, which reflects the viral logic to infect as much individuals as possible. Concluding, emerge of proteolytic sites in Omicron RBD, beneficial for proteasome cleavage, leading to release of public CD8+ T cell epitopes, might be one of the key factors that forced the SARS-CoV-2 to cross back the red line of the pandemic status.

## LIMITATIONS OF THE STUDY

ERAAP (ERAP1) cleaves broad spectrum of N-terminal amino acids in epitope precursors, but efficiency of the truncation may significantly differ [59]. Additionally, cytosolic aminopeptidases, acting before ERAAP, such as thimet oligopeptidase (TOP) and tripeptidyl peptidase II (TPPII) may contribute to the trimming of the HLA class I peptide ligands [60]. Thus, our *in silico* truncation of epitope precursors may not ideally reflects complexed intracellular pathway of antigen processing. Global correlative population analysis of frequency of human haplotypes and COVID-19 mortality in different countries potentially may yield either false-positive or false-negative result due to the rise of population immunity, different dynamic of vaccination, quarantine restrictions and regional healthcare organizational structure. Also, data on HLA haplotyping in some countries may be enriched in closed ethnic communities and thus be distort in terms of actual population coverage.

## SUPPLEMENTAL INFORMATION

Supplemental information includes Supplemental Tables 1-9 and Supplemental Data 1-2.

## Supporting information

Supplemental Information List

## Data Availability

Further information and requests for resources should be directed to and will be fulfilled by the lead contact, Alexey Belogurov. The mass spectrometry proteomics data have been deposited to the ProteomeXchange Consortium via the PRIDE partner repository with the dataset identifier PXD050265. The code and pipelines used for data analysis are available upon request. Raw data files are located at Mendeley Data (https://doi.org/10.17632/yc7ht4cgnc.1).

https://www.ebi.ac.uk/pride/archive/projects/PXD050265

## ACKNOWLEDGMENTS

This work was supported, in part, by RSF grants 21-74-10154 (to A.A.K.) (mass spectrometry and bioinformatic studies) and 23-74-00053 (to A.A.B. Jr.) (proteasome samples purification). The funders had no role in study design, data collection and interpretation, or the decision to submit the work for publication.

## AUTHOR CONTRIBUTIONS

Conceptualization, V.M.G. and A.A.B.Jr.; investigation, A.A.K., I.O.B., G.A.S., A.S.E., and Y.A.M.; resources, A.A.G., V.M.G., and A.A.B.Jr.; visualization, M.R., and A.A.B.Jr; validation, A.A.K. and A.A.B.Jr.; funding acquisition, A.A.K. and A.A.B.Jr.; project administration, A.A.K., I.V.S., D.S.M., and A.A.B.Jr.; supervision, A.A.K., I.V.S., D.S.M., and A.A.B.Jr.; writing – original draft, R.M., A.A.B., and A.A.B.Jr.; writing – review & editing, A.A.K., R.M., A.A.B., A.A.G., V.M.G., and A.A.B.Jr.

## MATERIALS AND METHODS

### Resource availability

#### Lead contact

Further information and requests for resources should be directed to and will be fulfilled by the lead contact, Alexey Belogurov.

#### Data and code availability

The mass spectrometry proteomics data have been deposited to the ProteomeXchange Consortium via the PRIDE [61] partner repository with the dataset identifier PXD050265. The code and pipelines used for data analysis are available upon request. Raw data files are located at Mendeley Data (https://doi.org/10.17632/yc7ht4cgnc.1).

#### Recombinant RBD proteins

cDNA coding for recombinant receptor binding domain (RBD) of the ancestorial SARS-CoV-2 strain (amino acids residues 330–528, Wuhan Hu-1) and its lineages Alpha B.1.1.7 (N501Y), Gamma P.1 (K417T, E484K, N501Y), Delta B.1.617.2 (L452R и T478K) and Omicron B.1.1.529 (G339D, S371L, S373P, S375F, K417N, N440K, G446S, S477N, T478K, E484A, Q493R,

G496S, Q498R, N501Y, Y505H) in frame with AviTag (GLNDIFEAQKIEWHE) and 6xHis tag were chemically synthesized (Evrogen, Russia) and cloned into the pFUSE vector using *EcoRI* and *NheI* restriction endonucleases. Recombinant proteins were produced by transient expression in HEK 293F cells, followed by purification on HiTrap Chelating column and size exclusion chromatography in PBS buffer on Superdex 75 10/300 GL according to the manufacturer’s protocols.

#### Mammalian Cells Constitutively Expressing HTBH-Tagged Proteasomes

HEK 293T (human embryonic kidney) and HeLa (human, cervical adenocarcinoma) cells were obtained from shared research facility "Vertebrate cell culture collection" of the Institute of Cytology Russian Academy of Sciences. All mammalian cell lines were cultured in DMEM medium (GIBCO, Thermo Fisher Scientific, Waltham, MA, USA). The media was supplemented with 10% fetal bovine serum (GIBCO, Thermo Fisher Scientific, Waltham, MA, USA) and 1% antibiotic-antimycotic (GIBCO, Thermo Fisher Scientific, Waltham, MA, USA). The cell lines were incubated in 37°C humidified incubator with 5% CO_2_. All cells were routinely tested for *Mycoplasma* contamination. The Sleeping beauty transposon system was used to generate HEK 293T and HeLa cells that overexpressed human PSMB4 proteasome subunit with HTBH tag. Cells were co-transfected with Sleeping Beauty transposon plasmid pSBi-Pur (PSMB4- HTBH, cDNA) and Sleeping Beauty transposase plasmid pCMV (CAT) T7-SB100 in ratio 10:1 with Lipofectamine 3000 (Thermo Fisher Scientific, Waltham, MA, USA). Three days after transfection, cells were maintained in selection medium (1 μg/mL puromycin in DMEM growth medium) for at least seven days. Sleeping Beauty transposon vector pSBbi-Pur (Addgene no.60523) and the pCMV(CAT)T7-SB100 (Addgene no. 34879) containing the cytomegalovirus (CMV) promoter and SB100X transposase were gifts to Addgene from Eric Kowarz [62] and Zsuzsanna Izsvak [63], respectively. The sequence encoding the PSMB4 proteasome subunit was amplified from cDNA isolated from HeLa cells via PCR, then overlapped with HTBH tag sequence and subcloned into the pSBbi-Pur vector.

#### Purification of HTBH-tagged 20S proteasomes

Stable HeLa or HEK293T cell lines expressing HTBH-tagged proteasomes were cultured until they reached 90% confluence, and then were washed with PBS buffer. The cell pellets were lysed using a buffer containing 30 mM Tris-HCl (pH 7.5), 5 mM MgCl_2_, 0.5% NP-40, 1 mM DTT, 1 mM PMSF. The lysates were centrifuged at 20,000g for 15 minutes to remove cellular debris, and the resulting supernatant was incubated with Streptavidin-agarose resin (Thermo Fisher Scientific, Waltham, MA, USA) overnight at 4°C with constant rotation. To purify the 20S proteasome, the streptavidin beads were washed twice with 10 volumes of the following buffer: 30 mM Tris-HCl (pH 7.5), 5 mM MgCl_2_, 1 mM TCEP, and 750 mM NaCl. Subsequently, they were washed another two times with same buffer without 750 mM NaCl. The beads were then resuspended in the required volume of wash buffer containing TEV protease, His (Genscript Biotech, Piscataway, NJ, USA), and were incubated at 30°C for 2 hours.

#### In-Gel Fluorescence with Me4BodipyFL-Ahx3Leu3VS Fluorescent Proteasome Probe

Equal amounts of 20S proteasomes were incubated with Me_4_BodipyFL-Ahx_3_Leu_3_VS for 30 min at 37°C in buffer containing 30 mM Tris-HCl pH 7.5, 1 mM TCEP. This was followed by adding sample buffer containing β-mercaptoethanol. Samples were analyzed using 15% sodium dodecyl sulfate polyacrylamide gel electrophoresis (SDS-PAGE). Wet gel slabs were imaged using the imaging system (ChemiDoc, Bio-Rad, Hercules, CA, USA) with appropriate filter settings (λ(ex/em) = 480/530 nm). Protein loading was verified by staining gels with a Coomassie blue stain.

#### Western blotting

Equal amounts of protein were mixed with 2x sample buffer containing β-mercaptoethanol and heated at 95 °C for 10 min. Proteins were separated by 15% SDS-PAGE and transferred to nitrocellulose membrane (Bio-Rad, Hercules, CA, USA), the membrane was then blocked with PBS-T containing 5% milk. All membranes were probed overnight with indicated primary antibodies in PBS-T with 0.5% milk at 4 °C (a-LMP2 (ab3328, abcam, UK), a-LMP7 (ab3329, Abcam, UK), a-LMP10 (ab183506, Abcam, UK), a-alpha subunits (BML-PW8195, Enzo Life Sciences, Inc, USA), a-PSMB5 (PA5-28086, Invitrogen, USA)), followed by 1 hour incubation with secondary antibodies in PBS-T with 0.5% milk (Goat Anti-Mouse IgG (Fc specific)–HRP (A2554, Sigma, USA), Goat Anti-Rabbit IgG-HRP (31460, Thermo Fisher Scientific, USA)). Bands were detected with ECL reagent (Bio-Rad, Hercules, CA, USA) using ChemiDoc imaging system (Bio-Rad, Hercules, CA, USA) and serial time exposure with signal saturation avoidance.

#### Analysis of peptidase activity of proteasomes

The peptidase activity of the proteasomes was assessed using a microplate reader (CLARIOstar plus, BMG Labtech, Ortenberg, Germany) at 37°C. A volume of 25 μL, consisting 0.125 μg of proteasome and 20 μM of either the fluorogenic substrates Suc-LLVY-AMC, Boc-LRR-AMC, Z-LLE-AMC or Ac-PAL-AMC, was used for the assay. The excitation wavelength was set at 380 nm, and the emission wavelength was set at 440 nm. The buffer used for measuring proteasome activity contained 30 mM Tris-HCl (pH 7.5), 5 mM MgCl_2_ and 1 mM TCEP.

#### RBD hydrolysis by purified proteasomes

The hydrolysis of RBD variants (1.5 μg) by 20S proteasomes (1 μg) was carried out in a 20 μL volume of buffer contained 20 mM Tris (pH 7.5), 5 mM MgCl_2_, and 1 mM DTT. The mixtures were incubated overnight at 37°C. Control RBD samples were incubated in same conditions without proteasomes.

#### LC-MS/MS Analysis

Totally we analyzed 50 samples, 10 samples per RBD: each RBD variant with either c20S or i20S (4 replicates, 8 samples totally) and control RBD samples (2 replicates). 1 uL of each sample was diluted with 10 uL of loading solution (5% v/v acetonitrile, 0.1% trifluoracetic acid) and 5 uL were injected. Injection was performed in trap-elute manner on trap column cartridge (PepMap Neo C_18_ 5 μM 300 μM x 5 mm, Thermo Scientific, USA) with 10 μL/min flow rate of loading solution. Peptides were separated on capillary column (Peaky, Reprosil Pur C_18_ AQ 1.9

μM, 75 μm x 30 cm, Molecta, Russia). Elution was performed with mobile phase gradient from 5% of solution B (80% acetonitrile, 0.1% formic acid) in solution A (0.1% formic acid) at flow rate of 250 nL/min to 50% of solvent B in 10 minutes. Detection was performed in data-dependent acquisition mode on high-resolution quadrupole-orbitrap tandem mass-spectrometer Exploris 480 (Thermo Scientific, USA). Electrospray ionization voltage was set at 2,200 volts. Precursor scan was performed for ions wit m/z from 350 to 1400 at resolution 60 000 (at 200 m/z). Up to 30 precursors with charge from 2 to 6 were subjected to fragment ion scan (resolution 15 000 at 200 m/z) with normalized collision energy set to 30%.

#### LC-MS Data Processing

Peptide identification was performed with FragPipe software. Search was performed against a database containing sequences of RBD variants, proteasome proteins and common contaminants. Non-specific cleavage was used, peptides with mass from 500 to 12000 kDa and length from 7 to 65 residues were considered. Mass tolerance was set to 20 ppm with further optimization. Oxidation of methionines, deamidation of asparagine and glutamine, N-terminal pyro-glutamine, N-term acetylation and serine, threonine and tyrosine phosphorylation formation were used as variable modifications. FDR levels were set to 1%, validation was performed with PeptideProphet and ProteinPropher. Label-free quantification was performed with MaxQuant software against a database of peptides identified with FragPipe with no cleavage at all with the same set of variable modifications. FDR was set at 1% for PSM and Protein levels. Label-free quantification and matching between runs were enabled. The mass spectrometry proteomics data are deposited to the ProteomeXchange Consortium via the PRIDE [61] partner repository with the dataset identifier PXD050265.

#### Bioinformatic processing

Identified peptides were filtered by mapping on corresponded RBD sequence (Wuhan-Hu-1 [www.ncbi.nlm.nih.gov/protein/1791269090], Alpha B.1.1.7, Delta B.1.617.2, Gamma B.1.1.28 (P.1), and Omicron B.1.1.529 (BA.1) [www.ecdc.europa.eu/en/covid-19/variants-concern]). Peptides, which overlapped with C-terminal AviTag-His, N-terminal non-RBD leader sequence or observed in samples without proteasomes were withdrawn. Resulted dataset contained 821 peptides (**Supplemental Table 1**). Normalization for each peptide was performed by dividing the values of its ion current by the total ion current (TIC) value across all observed RBD peptides. Final relative abundance of each peptide in RBD hydrolyzates represents average value from four independent replicates. Peptides ranging from 8 to 16 amino acids were recruited in analysis of human leukocyte antigen class I (HLA class I) binding. Specifically, we utilized the netMHCpan-4.1 algorithm [64] for 305 HLA class I alleles (**Supplemental Table 2**) covering 98% of the reqruited population (28,104 individuals with 4-digit HLA class I code or higher) as documented in the Allele Frequency Net Database [www.allelefrequencies.net/BrowseGenotype.aspx]. The database initially contains 82,130 samples, of which 28,104 samples were used with HLA class I data in a 4-digit code or higher for the alleles A, B, and C. Any alleles represented by a 6-digit code or higher were convoluted to a 4-digit code. Peptides with a length of 8 amino acids were directly inputted into the algorithm, while peptides with a length more than 8 amino acids were truncated to the length of 8, 9 and 10. For HLA class I alleles that occurred multiple times, the one with the lowest EL rank was selected. Only HLA variants with threshold lower or equal 0.5 were considered. For 237 peptides positive for HLA class I binding the average level of protection and percentage of individuals with at least one positive HLA class I allele (1-6) in each country were calculated (**Supplemental Table 3**). The CD8^+^ T cell epitopes sequences within RBD were extracted from IEDB database (**Supplemental Table 5**). The N-terminally extended proteasome-generated peptides were attributed the core CD8^+^ T cell epitopes (**Supplemental Table 6**) and further manually supplemented with data regarding positive CD8^+^ T cell assays (**Supplemental Table 7**). Amount of COVID-19 cases and related deaths were calculated from the file https://covid19.who.int/WHO-COVID-19-global-data.csv dated 20.12.2023 (**Supplemental Table 9**). Spearman correlation was calculated for each peptide between confirmed COVID-19- caused deaths and registered COVID-19 cases before or after December, 2021 (**Supplemental Table 7**). Analysis and figures were accomplished using a custom bash and R scripts available on request.

## SUPPLEMENTAL TABLES AND DATA LEGENDS

**Supplemental Table 1** (related to Figures 3, 4, 5). Global data on relative abundance of SARS-CoV-2 S protein RBD peptides in samples, generated by c20S and i20S proteasomes, calculated HLA class I EL ranks, percentage of HLA class I alleles coverage, average HLA class I haplotype protective index and percentage of individuals from 28,104 with at least 1 positive HLA class I allele in haplotype.

**Supplemental Table 2** (related to Figure 5). List of 305 HLA class I alleles, covering 98% haplotypes of 28,104 individuals deposited in Allele Frequency Net Database.

**Supplemental Table 3** (related to Figure 5). SARS-CoV-2 S protein RBD peptides (n=237) positive for HLA class I molecules binding and calculated average HLA class I haplotype protective index and percentage of individuals from 28,104 with at least 1 positive HLA class I allele in haplotype in different countries.

**Supplemental Table 4** (related to Figure 5E). Data on multiple Venn diagrams of S protein RBD peptides of SARS-CoV-2 strains generated by c20S and i20S proteasomes. Protective index (average amount of epitope-positive HLA class I alleles in world population human haplotypes, PI) and percentage of individuals from 28,104 with at least 1 positive HLA class I allele in haplotype for each cluster are listed.

**Supplemental Table 5** (related to Figure 6A). SARS-CoV-2 S protein RBD CD8+ T cell epitopes deposited in Immune Epitope Database (IEDB).

**Supplemental Table 6** (related to Figure 6A). Referencing of proteasome-generated N-terminally extended peptides to core HLA class I CD8+ T cell epitopes listed in the Immune Epitope Database (IEDB). Convoluted relative abundance of SARS-CoV-2 S protein RBD peptides in samples, generated by c20S and i20S proteasomes, Calculated HLA class I EL ranks, percentage of HLA class I alleles coverage, average HLA class I haplotype protective index and percentage of individuals from 28,104 with at least 1 positive HLA class I allele in haplotype.

**Supplemental Table 7** (related to Figure 6 and Figure 7B and 7D). IEDB core HLA class I CD8+ T cell epitopes, calculated HLA class I EL ranks, percentage of HLA class I alleles coverage, average HLA class I haplotype protective index and percentage of individuals from 28,104 with at least 1 positive HLA class I allele in haplotype, data on experimentally observed positive CD8^+^ T cell assays, convoluted relative abundance of SARS-CoV-2 S protein RBD peptides in samples and average HLA class I haplotype protective index and percentage of individuals from 28,104 with at least 1 positive HLA class I allele in haplotype in different countries. Spearman correlation between 496SFRPTYGVGH505 and 504GHQPYRVVVL513-positive HLA class I haplotypes in different populations and COVID-19-related deaths/cases ratio.

**Supplemental Table 8** (related to Figure 7C). Analysis of population frequency of SARS-CoV-2 S protein RBD 496SFRPTYGVGH505 and 504GHQPYRVVVL513-binding HLA class I molecules in different countries.

**Supplemental Table 9** (related to Figure 7B). Total amount of COVID-19-related deaths/cases in different countries before and after 12/01/2021.

**Supplemental Data 1** (related to Figure 5). Interactive plot representing relative abundance of SARS-CoV-2 S protein RBD peptides in samples, generated by c20S and i20S proteasomes with indication of positive HLA class I alleles and respective EL ranks.

**Supplemental Data 2** (related to Figure 5). Interactive plot with searchable interface representing sequence distribution of SARS-CoV-2 S protein RBD peptides, generated by proteasomes, with indication of positive HLA class I alleles and respective EL ranks.

